# Associations between rheumatoid arthritis clinical factors with synovial cell types and states

**DOI:** 10.1101/2023.04.27.23289104

**Authors:** Dana Weisenfeld, Fan Zhang, Laura Donlin, Anna Helena Jonsson, William Apruzzese, Debbie Campbell, Accelerating Medicines Partnership Program: Rheumatoid Arthritis Network, V. Michael Holers, Ellen Gravallese, Larry Moreland, Susan Goodman, Michael Brenner, Soumya Raychaudhuri, Andrew Filer, Jennifer Anolik, Vivian Bykerk, Katherine P. Liao

**Affiliations:** Division of Rheumatology, Inflammation, and Immunity, Brigham and Women’s Hospital and Harvard Medical School, Boston, MA, USA; Division of Rheumatology, University of Colorado School of Medicine, Aurora, CO, USA; Center for Health Artificial Intelligence, University of Colorado School of Medicine, Aurora, CO, USA; Division of Rheumatology, Hospital for Special Surgery, New York, NY, USA; Weill Cornell Medicine, New York, NY, USA; Accelerating Medicines Partnership® Program: Rheumatoid Arthritis and Systemic Lupus Erythematosus (AMP® RA/SLE) Network; Division of Allergy, Immunology and Rheumatology, University of Rochester Medical Center, Rochester, NY, USA; Division of Rheumatology and Clinical Immunology, University of Pittsburgh School of Medicine, Pittsburgh, PA, USA; Center for Data Sciences, Brigham and Women’s Hospital, Boston, MA, USA; Division of Genetics, Department of Medicine, Brigham and Women’s Hospital and Harvard Medical School, Boston, MA, USA; Department of Biomedical Informatics, Harvard Medical School, Boston, MA, USA; Broad Institute of MIT and Harvard, Cambridge, MA, USA; Rheumatology Research Group, Institute for Inflammation and Ageing, NIHR Birmingham Biomedical Research Center and Clinical Research Facility, University of Birmingham, Queen Elizabeth Hospital, Birmingham, UK; MRC Versus Arthritis Centre for Musculoskeletal Ageing Research and the Research into Inflammatory Arthritis Centre Versus Arthritis, University of Birmingham, Birmingham, UK

## Abstract

**Objective:** Recent studies have uncovered diverse cell types/states in the RA synovium; however limited data exist correlating these findings with patient-level clinical information. Using the largest cohort to date with clinical and multi-cell data, we determined associations between RA clinical factors with cell types/states in the RA synovium.

**Methods:** The Accelerated Medicines Partnership Rheumatoid Arthritis study recruited subjects with active RA on no DMARDs or inadequate response to methotrexate (MTX) or tumor necrosis factor inhibitors. RA clinical factors were systematically collected. Biopsies were performed on an inflamed joint and tissue disaggregated and processed with a CITE-seq pipeline from which cell type percentages and cell type abundance phenotypes (CTAP) were derived: endothelial/fibroblast/myeloid (EFM), fibroblasts (F), myeloid (M), T/B cells (TB), T cells/fibroblasts (TF), T/myeloid cells (TM). Correlations were measured between RA clinical factors, % cell type, and CTAPs.

**Results:** We studied 72 subjects, mean age 57 years, 75% female, 83% seropositive, mean RA duration 6.6 years, mean DAS28-CRP3 4.8. Higher DAS28-CRP3 correlated with higher % T cells (p<0.01). Subjects on MTX not on bDMARD had higher %B cells vs no DMARDs (p<0.01). The majority of subjects on bDMARDs were categorized as EFM (57%), while none were TF. No significant difference was observed across CTAPs for age, sex, RA disease duration, DAS28-CRP3.

**Conclusion:** In this comprehensive screen of clinical factors, we observed differential associations between DMARDs and cell phenotypes, suggesting that RA therapies, more than other clinical factors may impact cell type/state in the synovium and ultimately response to subsequent therapies.

## INTRODUCTION

Rheumatoid arthritis (RA) is the most common autoimmune inflammatory joint disease worldwide, typically presenting as symmetric small joint polyarthritis (1,2). While there is consensus regarding diagnostic and classification criteria for RA (3), multiple underlying pathways lead to a clinical presentation of RA. Recent studies have characterized the diverse cell populations in the RA synovium incorporating histology in combination with gene expression, single-cell (scRNA-seq) data or cytometry (4–7). Additionally, the incorporation of multimodal CITE-seq to histology and RNA-seq data have enabled the creation of a comprehensive synovial cell atlas of cell types and states (8). Due to the novelty of these approaches, as well as the infrastructure required to collect patient level data, there is limited information on the associations between patient-level clinical measurements with cell types and states.

Understanding these diverse pathways through detailed studies of both the cell types and expression have demonstrated promise in guiding rheumatologists in identifying the optimal targeted therapy from the many options for a particular patient (9–11). Further integrating patient-level clinical data can inform whether cellular states in the synovium correlate with the overall clinical picture. Thus, the objective of this study was to determine the correlations between cell types and states with RA clinical factors at the patient level.

## METHODS

This study was performed using data from the Accelerated Medicine Partnership (AMP) RA Network. Briefly, patients were recruited from 13 centers with inclusion criteria age≥18, fulfilled the 1987 or 2010 ACR/EULAR RA Classification criteria with ≥2 tender and swollen joints and a CDAI≥10 (8). In addition, recruitment focused on subjects with active RA in one of 3 categories: (1) not yet commenced disease modifying anti-rheumatic drugs (DMARDs), (2) on methotrexate (MTX) and not on a biologic (bDMARD), or (3) on a tumor necrosis factor inhibitor (TNFi). The study was performed in accordance with protocols approved by the institutional review board.

### RA clinical factors

RA clinical factors were assessed at the baseline visit including age, sex, RA disease duration, rheumatoid factor (RF) and anti-cyclic citrullinated peptide (anti-CCP) antibody positivity, high sensitivity C-reactive protein (hsCRP) levels, clinical disease activity score (CDAI), the disease activity score 28-CRP3 (DAS28-CRP3), as well as the components of the risk scores. Other assessments included physical function using the health assessment questionnaire (HAQ) and smoking status. Exploratory analyses were also performed to examine potential differences across treatment. We categorized subjects based on their baseline treatments: (a) not on a DMARD, (b) MTX and not on a b/tsDMARD; and (c) any bDMARD or janus kinase inhibitor (JAKi), category referred to as b/tsDMARD. Subjects in category (b) were allowed to be on combination therapy with another non-biologic (nb) DMARD. For certain analyses, category (b) was expanded to include subjects on any nbDMARD but no b/tsDMARD.

### Synovial cellular phenotype

Six major cell types were observed in the synovial tissue samples: T, B and plasma, natural killer (NK), myeloid, stromal, and endothelial cells. In this study, we characterized the synovial cellular data as: (1) % of each cell type in synovial tissue, and (2) cell type abundance phenotypes (CTAPs). The % cell type was calculated by dividing the total number of cells for each cell type by the total number of cells extracted from the disaggregated tissue. Methods for stratifying tissues into CTAPs were previously described (8). Briefly, the CTAPs were identified based on hierarchical clustering on the cell-type abundance, which characterizes the heterogeneity of RA synovial tissues. The 6 CTAPs for this study, named based on the most abundant cell type were: (1) endothelial, fibroblast, and myeloid cells (EFM), (2) fibroblasts (F), (3) myeloid (M), (4) T and B cells (TB), (5) T cells and fibroblasts (TF), (6) T and myeloid cells (TM). In short, CTAPs offer a classification of tissue heterogeneity based on cell frequency, which can be generalized to multiple technologies, with a goal to inform further clinical association studies.

We performed correlations between key RA clinical data and cell type % using Kendall’s tau for continuous variables and point bi-seral correlation for binary variables. Fisher’s exact test or ANOVA were performed to test differences in proportions or means, respectively, across CTAP categories. In this discovery cohort, we considered associations p<0.01 as significant, but also reported p<0.05. All univariate correlations with p<0.01 were followed by linear regression analysis adjusting for age, sex, and RA duration.

## RESULTS

In total, n=72 subjects had clinical and synovial cell data with mean age 57 years, 75% female, 71% White, 18% Black, 28% Hispanic; 83% seropositive, mean RA duration 6.6 years, DAS28-CRP3 4.8, and hsCRP 19.6mg/L (**Table 1**).

**Table 1.** Comparison of demographics and RA clinical factors across cell type abundance phenotypes (CTAP).

| Clinical characteristics | Overall | CTAP |  |  |  |  |  | p-value |
| --- | --- | --- | --- | --- | --- | --- | --- | --- |
|  |  | EFM | F | M | T+B | T+F | T+M |  |
| N | 72 | 7 | 11 | 18 | 14 | 8 | 12 |  |
| Age, yrs | 56.5 ±15.1 | 66.6 ±7.11 | 56.2 ±14.4 | 56.2 ±21.8 | 54.5 ±12.9 | 58.0 ±9.07 | 51.1 ±14.4 | 0.435 |
| Female, N (%) | 54 (75.0) | 6 (85.7) | 7 (63.6) | 13 (72.2) | 11 (78.6) | 6 (75.0) | 9 (75.0) | 0.963 |
| RA duration, median yrs [IQR] | 2.05<br>[0.10, 9.67] | 15.4<br>[4.90, 18.1] | 0.66<br>[0.12, 5.53] | 1.95<br>[0.04, 9.39] | 3.93<br>[0.47, 12.5] | 2.71<br>[0.36, 7.38] | 0.47<br>[0.02, 4.22] | 0.268 |
| Anti-CCP+, N (%) | 54 (76.1) | 6 (85.7) | 9 (81.8) | 9 (50.0) | 13 (92.9) | 7 (100.0) | 10 (83.3) | 0.041 |
| DAS28-CRP3 | 4.84 ±1.39 | 4.47 ±0.71 | 4.80 ±1.46 | 4.61 ±1.34 | 4.81 ±1.34 | 5.34 ±2.56 | 5.43 ±1.49 | 0.626 |
| hsCRP, mg/L, median [IQR] | 8.40<br>[5.00, 21.0] | 6.00<br>[3.50, 7.00] | 7.00<br>[3.00, 12.0] | 9.46<br>[4.40, 20.5] | 10.0<br>[7.00, 37.1] | 29.0<br>[18.0, 61.2] | 8.25<br>[5.03, 34.5] | 0.351 |
| **RA clinical factors differing across CTAPs** |  |  |  |  |  |  |  |  |
| HAQ* | 1.27 ±0.65 | 1.20 ±0.55 | 1.08 ±0.55 | 1.21 ±0.62 | 1.07 ±0.47 | 1.91 ±0.76 | 1.46 ±0.74 | 0.041 |
| Patient global** | 5.75 ±2.25 | 4.83 ±1.49 | 4.82 ±1.65 | 6.03 ±2.58 | 5.12 ±2.78 | 7.75 ±1.56 | 6.55 ±1.12 | 0.031 |
| **RA treatments, N (%)** |  |  |  |  |  |  |  |  |
| No DMARD | 29 (40.3) | 1 (14.3) | 4 (36.4) | 11 (61.1) | 5 (35.7) | 4 (50.0) | 4 (33.3) | 0.343 |
| MTX, no b/tsDMARD*** | 23 (31.9) | 0 (0.0) | 4 (36.4) | 5 (27.8) | 6 (42.9) | 3 (37.5) | 4 (33.3) | 0.466 |
| Any b/tsDMARD | 13 (18.1) | 4 (57.1) | 1 (9.1) | 1 (5.6) | 3 (21.4) | 0 (0.0) | 3 (25.0) | 0.036 |
| Any DMARD | 43 (59.7) | 6 (85.7) | 7 (63.6) | 7 (38.9) | 9 (64.3) | 4 (50.0) | 8 (66.7) | 0.343 |

Mean ±SD shown unless otherwise specified. The overall column includes two subjects who were not classified into CTAPs. Fischer's exact test used to test for differences across CTAPs for binary variables; One-way ANOVA used for continuous variables. EFM=endothelial, fibroblast, myeloid; F=fibroblast; M=myeloid; T+B= T and B cells; T+F= T cells and fibroblasts; T+M= T and myeloid cells. \*HAQ, Health Assessment Questionnaire: 0=no difficulty through 3=unable to do activities. \*\*Patient global: 0=no RA activity through 10=worst RA activity. \*\*\*Can be in combination with nbDMARDs.

While there was considerable heterogeneity among samples, the majority of synovial tissue was populated by T cells (30%), followed by fibroblasts (25%), and myeloid cells (24%) (**Figure 1**). Both anti-CCP and RF positivity correlated with lower % myeloid cells, and RF+ was correlated with higher % T cells; anti-CCP had a similar correlation (**Figure 2**). Higher DAS28-CRP3 was correlated with higher % T cells in the synovium. This association remained about adjusting for age, sex, and RA disease duration.

**Figure 1.**
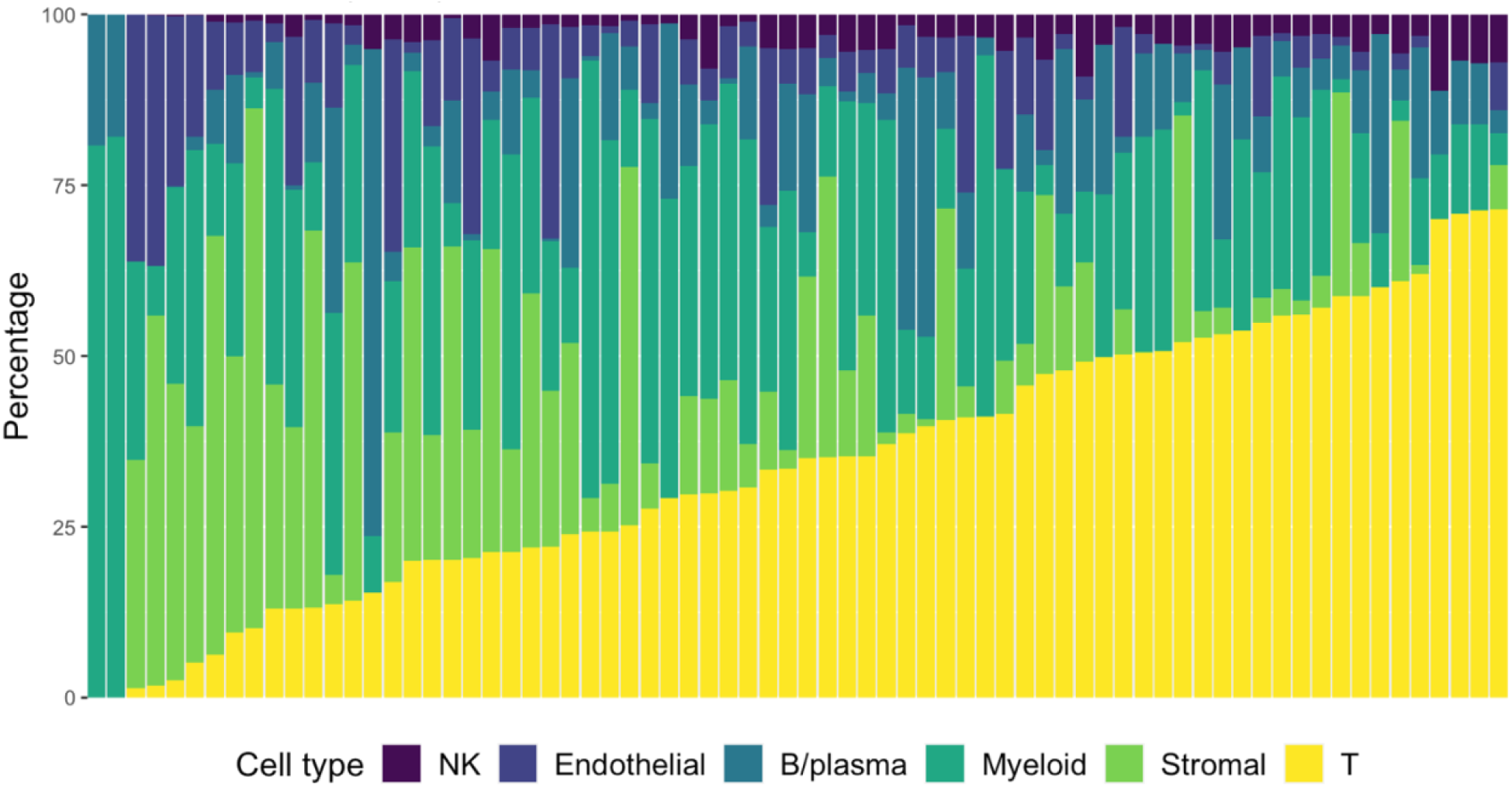
Percentage of T, B and plasma, natural killer (NK), myeloid, stromal, and endothelial cells in the RA synovial tissue with each bar representing an individual subject.

**Figure 2.**
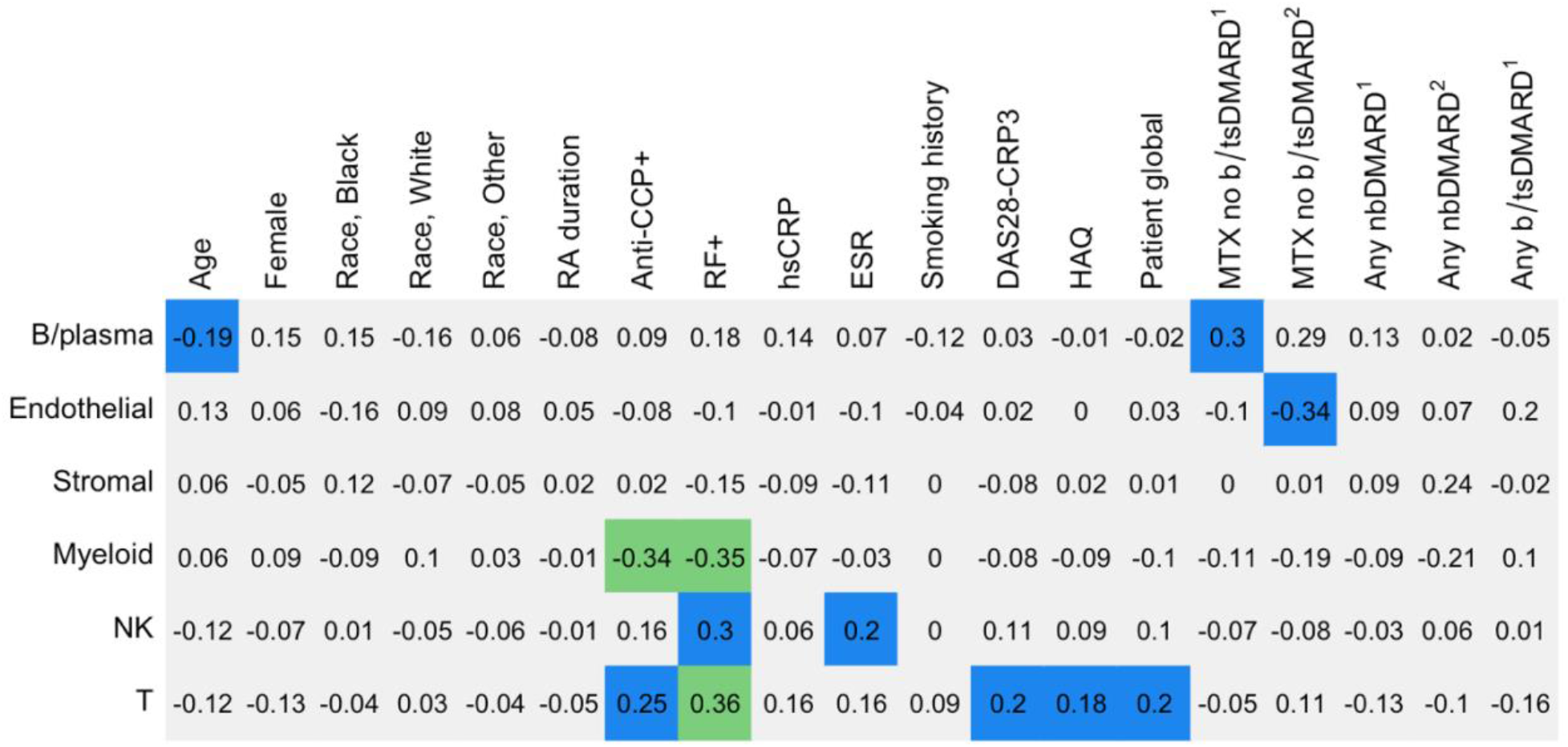
Correlations between cell type % with clinical factors; p-value<0.01 in green, p≥0.01 and <0.05 in blue. Pearson’s rho shown for binary variables; Kendall’s tau shown for continuous variables. ^1^Reference group is subjects not on a DMARD. ^2^Reference group is subjects on a bDMARD.

When examining variation in % cell type across baseline treatments, subjects on MTX but no b/tsDMARDs had higher %B cells compared to subjects on no DMARDs. Compared to subjects on a b/tsDMARD, subjects on MTX had a lower % endothelial cells. Both associations were independent of age, sex, and disease duration. There was no difference in mean DAS28-CRP3 across treatment groups, p=0.55 (**Supplementary Table 1**). Of note, 30% of subjects on MTX but no b/tsDMARD were on combination therapy with another nbDMARD.

Among the CTAPs, the largest proportion of subjects were classified in the M group (26%), followed by TB (20%), and TM (17%). Patients with tissue classified as TF had the highest HAQ and patient global scores, corresponding with more difficulty performing activities of daily living and higher patient reported RA disease activity (**Table 1**). The majority of subjects in EFM were on any DMARD. More specifically, among those on a DMARD, the majority of b/tsDMARDs were categorized in the EFM CTAP (57%). In contrast, while 50% of subjects in TF were on any DMARD, none were on a b/tsDMARD. Furthermore, among subjects categorized in the EFM CTAP, none were on MTX with no bDMARD. No significant difference was observed across CTAPs for age, sex, RA disease duration, DAS28-CRP3, and hsCRP.

## DISCUSSION

In this study, we identified correlations between RA clinical factors with synovial cell types and states using the largest resource to date with these integrated data. In line with prior studies, T cells were the most common cell-type in RA synovial tissue and correlated with higher RA disease activity (2,12,13). Using novel data incorporating RNA-seq, and CITE-seq data to derive CTAPs, we observed that the proportion of subjects in RA treatments groups differed across CTAPs, despite no significant differences in age, sex, RA disease duration or disease activity. Overall, our data suggest that RA therapies may impact the cellular phenotype in the synovium.

Individuals with active RA on MTX as monotherapy or in combination with another nbDMARD but not on a b/tsDMARD (MTX, no b/tsDMARD) were more likely to have a higher %B-lymphocyte compared to subjects on no DMARDs. These findings corroborate a separate study identifying a B-lymphocyte rich subset in both nbDMARD and bDMARD inadequate responders (10). In that study, subjects with low B cell expression in the synovial tissue were more likely to respond to tocilizumab compared to rituximab which targets B cells. In the present study, subjects on b/tsDMARDs were more likely to belong in the lymphocyte poor EFM CTAP. Additionally, the profile of EFM tissue was distinct from osteoarthritis tissue and thus EFM tissue likely do not represent an immunologically quiescent subset (8). Other aspects of this group included the fact that 0% of MTX, no bDMARD subjects were categorized as EFM, raising the possibility that treatment may influence CTAP. We observed no significant differences in demographics, disease duration or disease activity across CTAP categories to explain why subjects on certain treatments were enriched in some CTAPs and not others. Further longitudinal studies are needed to disentangle whether the EFM phenotype is a result of longer disease duration, a change that may be associated with bDMARD therapy, particularly TNFi therapy, or a combination of both, and ultimately whether a specific targeted therapy is more effective for this subgroup.

Patient reported outcomes (PROs), specifically physical function as measured by the HAQ and the patient global, differed across CTAPs. Subjects with synovial tissue classified as TF reported the most limitations in their physical function and highest RA activity at the time of the biopsy. Among subjects classified none of the subjects were on a b/tsDMARD at the time. Together, these results raise the possibility that the TF CTAP correlates with a group of RA subjects who are earlier in their treatment course for RA, and at a period where their symptoms impact their physical function. In this study, RA disease duration alone was unable to account for the differences in correlation for physical function or treatments across CTAPs. The connection between immune-cell transcriptional profiles and patient reported symptoms has been observed in a prior study using blood rather than synovial tissue. In a longitudinal study patient reported flares were correlated with increased activation of naïve B cells as measured by RNA-seq (14). Overall, these findings suggest that CTAPs may be characterized at the patient level by patient symptoms as reported by PROs and should be considered in future and ongoing studies of immunophenotyping.

A goal of this study was to serve as a resource reporting summary data from a novel discovery cohort. However, a limitation is that there are not yet sufficiently sized cohorts for replication. Additionally, while we studied the largest cohort with integrated clinical and cellular phenotyping to date, our study remains limited by sample size and thus limited power to detect associations.

Future directions include longitudinal studies with repeat biopsies which will provide key information on whether cell type and state change with treatment and ultimately, whether baseline cellular phenotypes can predict response to therapy. Overall, the correlations identified in this study suggest that RA treatments, more than RA duration and other clinical factors impact cellular phenotypes in the synovium. Data on cell percentages in the synovium and CTAP classification across treatment groups linked with RA clinical characteristics can be used to inform and power the design of future clinical trials as we continue to advance toward precision medicine in RA.

## Data Availability

All raw and processed data will be available upon publication of the primary methodology paper for the study, https://doi.org/10.1101/2022.02.25.481990.

## Accelerating Medicines Partnership Program: Rheumatoid Arthritis and Systemic Lupus Erythematosus (AMP RA/SLE) Network includes

Jennifer Albrecht^7^, Jennifer L. Barnas^7^, Joan M. Bathon^15^, Ami Ben-Artzi^16^, Brendan F. Boyce^17^, David L. Boyle^18^, S. Louis Bridges Jr.^5,19^, Hayley L. Carr^13^, Arnold Ceponis^18^, Adam Chicoine^1^, Andrew Cordle^20^, Michelle Curtis^1,9–12^, Kevin D. Deane^2^, Edward DiCarlo^21^, Patrick Dunn^22,23^, Gary S. Firestein^18^, Lindsy Forbess^13^, Laura Geraldino-Pardilla^15^, Peter K. Gregersen^24^, Joel M. Guthridge^25^, Maria Gutierrez-Arcelus^1,9–12,26^, Siddarth Gurajala^1,9–12^, V. Diane Horowitz^24^, Laura B. Hughes^27^, Kazuyoshi Ishigaki^1,9– 12,28^, Lionel B. Ivashkiv^5,19^, Judith A. James^25^, Joyce B. Kang^1,9–12^, Gregory Keras^1^, Ilya Korsunsky^1,9–12^, Amit Lakhanpal^5,19^, James A. Lederer^29^, Zhihan J. Li^1^, Yuhong Li^1^, Arthur M. Mandelin II^30^, Ian Mantel^5,19^, Mark Maybury^13^, Andrew McDavid^31^, Mandy J. McGeachy^8^, Joseph Mears^1,9,11,12,29^, Nida Meednu^7^, Nghia Millard^1,9–12^, Aparna Nathan^1,9–12^, Alessandra Nerviani^32^, Dana E. Orange^19,33^, Harris Perlman^30^, Costantino Pitzalis^32^, Javier Rangel-Moreno^7^, Deepak A. Rao^1^, Karim Raza^13^, Yakir Reshef^1,9–12^, Christopher Ritchlinro^7^, Felice Rivellese^32^, William H. Robinson^34^, Laurie Rumker^1,9–12^, Ilfita Sahbudi^13^, Saori Sakaue^1,9–12^, Jennifer A. Seifert^2^, Kamil Slowikowski^11,12,35,36^, Melanie H. Smith^19^, Darren Tabechian^7^, Dagmar Scheel-Toellner^13^, Paul J. Utz^34^, Gerald F. M. Watts^1^, Kevin Wei^1^, Kathryn Weinand^1,9–12^, Michael H. Weisman^16,34^, Qian Xiao^1,9–12^, Zhu Zhu^1^

^15^ Division of Rheumatology, Columbia University College of Physicians and Surgeons, New York, NY, USA.

^16^ Division of Rheumatology, Cedars-Sinai Medical Center, Los Angeles, CA, USA.

^17^ Department of Pathology and Laboratory Medicine, University of Rochester Medical Center, Rochester, NY, USA.

^18^ Division of Rheumatology, Allergy and Immunology, University of California, San Diego, La Jolla, CA, USA.

^19^ Hospital for Special Surgery, New York, NY, USA.

^20^ Department of Radiology, University of Pittsburgh Medical Center, Pittsburgh, PA, USA.

^21^ Department of Pathology and Laboratory Medicine, Hospital for Special Surgery; New York, NY, USA.

^22^ Division of Allergy, Immunology, and Transplantation, National Institute of Allergy and Infectious Diseases, National Institutes of Health, Bethesda, MD, USA.

^23^ Northrop Grumman Health Solutions, Rockville, MD, USA.

^24^ Feinstein Institute for Medical Research, Northwell Health, Manhasset, New York, NY, USA.

^25^ Department of Arthritis & Clinical Immunology, Oklahoma Medical Research Foundation, Oklahoma City, OK, USA.

^26^ Division of Immunology, Department of Pediatrics, Boston Children’s Hospital and Harvard Medical School, Boston, MA. US.

^27^ Division of Clinical Immunology and Rheumatology, Department of Medicine, University of Alabama at Birmingham, Birmingham, AL, USA.

^28^ Laboratory for Human Immunogenetics, RIKEN Center for Integrative Medical Sciences, Yokohama, Japan.

^29^ Department of Surgery, Brigham and Women’s Hospital and Harvard Medical School, Boston, MA, USA.

^30^ Division of Rheumatology, Department of Medicine, Northwestern University Feinberg School of Medicine, Chicago, IL, USA.

^31^ Department of Biostatistics and Computational Biology, University of Rochester School of Medicine and Dentistry; Rochester, NY, USA.

^32^ Centre for Experimental Medicine & Rheumatology, William Harvey Research Institute, Queen Mary University of London; London, UK.

^33^ Laboratory of Molecular Neuro-Oncology, The Rockefeller University, New York, NY, USA.

^34^ Division of Immunology and Rheumatology, Institute for Immunity, Transplantation and Infection, Stanford University School of Medicine, Stanford, CA, USA.

^35^ Center for Immunology and Inflammatory Diseases, Department of Medicine, Massachusetts General Hospital (MGH), Boston, MA, USA.

^36^ MGH Cancer Center, Boston, MA, USA.

