## Supplementary Table 1 for "Associations between rheumatoid arthritis clinical factors with synovial cell types and states"

**Supplementary Table 1.** Comparison of clinical characteristics across baseline RA treatment categories.

| | **(a) No DMARD** | **(b) nbDMARD** | **(c) b/tsDMARD** | **p-value** |
|---|---|---|---|---|
| N | 29 | 30 | 13 | |
| Demographics | | | | |
| Age, years | 56.6 ±15.4 | 54.1 ±16.8 | 61.7 ±7.92 | 0.323 |
| Female, N (%) | 18 (62.1) | 25 (83.3) | 11 (84.6) | 0.136 |
| Race, N (%) | | | | |
| White | 19 (65.5) | 20 (66.7) | 12 (92.3) | 0.173 |
| Black | 5 (17.2) | 7 (23.3) | 1 (7.7) | 0.518 |
| Other | 5 (17.2) | 1 (3.3) | 0 (0.0) | 0.122 |
| RA factors | | | | |
| RA duration, median years [IQR] | 0.05 [0.01, 1.47] | 6.77 [1.89, 11.6] | 5.47 [4.07, 16.0] | <0.001 |
| Seropositive, N (%) | 24 (82.8) | 24 (82.8) | 11 (84.6) | 1.000 |
| CCP positive, N (%) | 21 (72.4) | 22 (75.9) | 11 (84.6) | 0.718 |
| RF positive, N (%) | 22 (75.9) | 20 (71.4) | 8 (61.5) | 0.651 |
| RA clinical measures | | | | |
| hsCRP, mg/L median [IQR] | 8.50 [5.50, 18.5] | 10.0 [6.00, 24.0] | 6.00 [3.00, 12.0] | 0.466 |
| ESR, mm/hr | 44.1 ±30.8 | 38.7 ±29.1 | 25.5 ±19.3 | 0.176 |
| Smoking history, N (%) | 16 (55.2) | 15 (50.0) | 7 (53.8) | 0.951 |
| DAS28-CRP3 | 4.69 ±1.20 | 4.94 ±1.63 | 4.96 ±1.32 | 0.774 |
| HAQ | 1.34 ±0.73 | 1.35 ±0.62 | 0.95 ±0.41 | 0.135 |

Mean ±SD shown unless otherwise noted. Fischer’s exact test used to test for differences across CTAPs for binary variables; One-way ANOVA used for continuous variables.
(a) Not on a DMARD at baseline visit; (b) Taking one or more nbDMARDs, not on a concurrent b/tsDMARD; (c) On a b/tsDMARD and could be on a concurrent nbDMARD.
